# Extracting clinical chief complaints from patient-physician conversations with a symbolic reasoning model

**DOI:** 10.64898/2025.12.10.25341937

**Authors:** Vivek Kanpa, David D’Onofrio, Nandini Samanta, Mikio Tada, Shrey Patel

**Author notes:** Corresponding Author: Vivek Kanpa, Postal Address: 50 E 98 Street, New York, NY 10029. These authors contributed equally to this work (Vivek Kanpa, David D’Onofrio).

## Abstract

Generative artificial intelligence (GenAI) applications have been at the forefront of clinical documentation assistants, aiming to reduce physician notetaking burden. However, GenAI systems are resource-intensive, and deployment in low-resource healthcare settings can be challenging and cost prohibitive. We present a symbolic reasoning model (SRM) for detecting chief complaints from clinical conversations and evaluate it against two large language models (LLMs), Gemma2-9b and Llama3.3-70B-Versatile. We use a benchmarking dataset of simulated doctor-patient conversations to develop an SRM leveraging proximity-based keywords and Zipf’s Law to extract complaints. Three physicians from different hospital systems evaluate the accuracy and clinical utility of the LLMs and SRM. The SRM achieves comparable or superior performance to LLMs across three conversational modalities of varying complexity (cross-modality mean utility score: 5.77 ± 0.78 out of 10). Runtime analyses demonstrated that the SRM executes substantially faster than LLMs, though with higher local RAM usage, contrasted with the cloud-based execution of LLMs. Conversational adjustments by physicians to acknowledge a third-party scribe system substantially improved utility of SRM-detected complaints. These findings support SRM integration as a plausible solution for automating documentation in under-resourced healthcare environments.

## Background and Significance

The use of GenAI in clinical settings can recapitulate physician notes and patient summaries, reducing the manual workload by healthcare workers to summarize physician notes^[1,2]^. The deployment of GenAI in healthcare organizations necessitate structures that ensure accuracy, explainability and minimizing bias even as data and practices evolve over time^[3]^. Automating the scribing process has the potential to mitigate notetaking demands on physicians while lowering medical documentation costs, staying in line with the quadruple aim of healthcare^[4]^. However, this supporting infrastructure is resource-intensive and infeasible to implement in many resource-poor healthcare settings. Implementing virtual scribing software creates substantial barriers to equitable adoption, disproportionately benefiting providers in high-resource environments^[5]^. Explicit rules-based algorithms, which require less computing power, modest infrastructure, and less technical expertise, represent a feasible and effective alternative that can significantly reduce these disparities by enabling implementation even in resource-limited settings^[6]^.

### Objectives

Our work aims to evaluate the potential of explicit rules-based algorithms for computer-aided clinical scribing as an alternative to resource-intensive GenAI. The objective of this investigation is to compare Gemma2-9b, Llama3.3-70B-Versatile, and a clinician-aware symbolic reasoning model powered by Zipf’s law and keyword detection to summarize chief complaints from follow-up and routine visits. We show that the rules-based SRM model can achieve performance on par with GenAI without being resource-intensive.

## Methods

### Data Source

Patient-physician conversations were sourced from the benchmarking dataset in Yim et al., 2023^[7]^. There are 87 simulated medical conversations recorded by a speech-to-text tool between a patient actor and certified physician in an outpatient care setting. The dataset was subdivided into 67 training conversations and 20 test set conversations. Each dialogue is accompanied by a ground truth chief complaint which was determined by physicians.

The dialogues took one of three conversational modalities, which are referred to as virtual assistant (virtassist), virtual scribe (virtscribe), and ambient clinical intelligence (aci). Dialogues from virtscribe and virtassist both include acknowledging the presence of a notetaker, either human or computational (i.e., Dragon Medical Virtual Assistant). Aci differs from both in that it does not require the clinician to modify language or acknowledge a third party. These modalities are used to simulate the conversation mannerisms a physician might use when addressing a patient, in the presence or absence of a digital assistant or human scribe, to evaluate the performance of various LLMs in all conversational situations.

### Detecting chief complaints

We developed a novel symbolic reasoning model (SRM) and compared it against existing LLMs to detect chief complaints from within the dialogue.

First, we tokenize conversations and score the frequency of words used by physicians in training set dialogues. Then, we selected those phrases which were proximal (within the same sentence) to the ground truth chief complaint (i.e “presents with”), hereafter referred to as flags. By parsing each conversation using these flags, we generated reading frames to detect candidate chief complaints directly from the text. The reading frame begins at the keyword and extends until the end of the physician’s sentence. Zipf’s Law was used to transform each token in the reading frame to a numeric value representing the logarithm of the token’s occurrence per million words in the English language. Reading frames were pruned where the change in Zipf’s score between tokens exceeds 1.0 (representing a tenfold change in occurrence).

The existing LLM models, Gemma2-9b-it and Llama3.3-70B-Versatile, were prompted to detect the most likely chief complaint from the patient in the conversation provided^[8,9]^. A second set of these models were asked to do this same task, but prompted to detect the chief complaint as succinctly as possible. We evaluated the zero-shot capability of Gemma2-9b-it and Llama3.3-70B-Versatile on the test data because GenAI may not be fine-tuned at each hospital in a real deployment setting. The code for each of the five models is available at https://github.com/david-donofrio/datathon.

### Evaluation metrics

We ask a team of three outpatient physicians to independently score the accuracy and clinical utility of each detected chief complaint, given the ground truth chief complaint. Physicians were blind to the model that produced each detected chief complaint. The 100 detection-ground truth pairs (5 models predicting 20 test set conversations) were randomly shuffled to reduce the effect of reviewer burnout bias and fluctuations in scoring judgement.

The SRM model was run 200 times and each LLM model was run 30 times to collect usage metrics such as total time (ms), maximum RAM memory (MB), and token usage/cost to predict across the entire test set. Significance across models was tested using Welch’s t-tests.

## Results

The utility scores of detected chief complaints ascribed by the three physician reviewers generally demonstrate high concordance (Supplementary Figure 1). Accuracy scoring also aligns between reviewers well; in 56% of detected chief complaints, the three physicians unanimously agreed on an accuracy grade. In the remaining 44 cases, SK classified all detections as inaccurate, while EC and PD classified 30 of these detections as accurate. A table of all model chief complaint detections, with respect to the ground truth for each conversation, is made available in the supplementary data (Supplementary Table 1).

Virtassist conversations enabled chief complaint detections with the highest average utility score across all five detection models (5.053 +/– 1.341). Aci conversations followed closely behind (4.839 +/– 1.276) while virtscribe conversations generate an average cross-model utility score a full point lower (3.933 +/– 2.038). Averaged across conversation modalities, the SRM achieves the highest scoring utility by physician reviewers (5.770 +/– 0.777), followed by the two Llama models, and then the two Gemma models. While Llama3.3-70B-Versatile generates higher cross-modality average utility detections when prompted for succinctness than without the succinctness prompt, Gemma2-9b seemingly performs worse when prompted for succinctness. Gemma2-9b consistently performs the worst within each conversation modality, while the SRM gloats best-in-class performance in virtscribe and virtassist tasks. In aci conversations, the SRM is bested by LLM alternatives, ranking 4th out of the 5 models (Figure 1 & Table 1).

**Figure 1.**
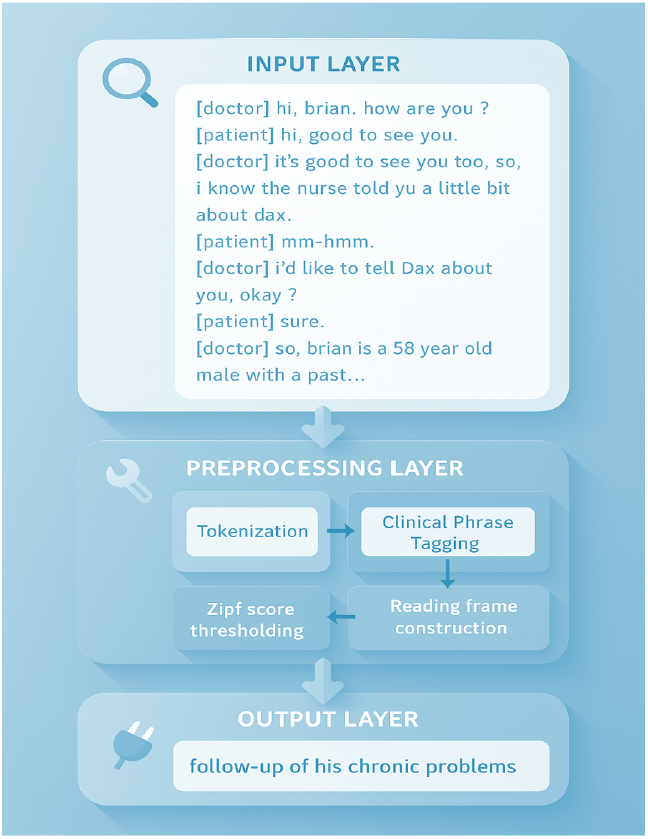
An example of the SRM framework. A conversation modality is provided to the model as an input layer and separated into tokens that are given a Zipf score. The output provided is a set of key words with the highest between-word Zipf score change.

**Figure 2.**
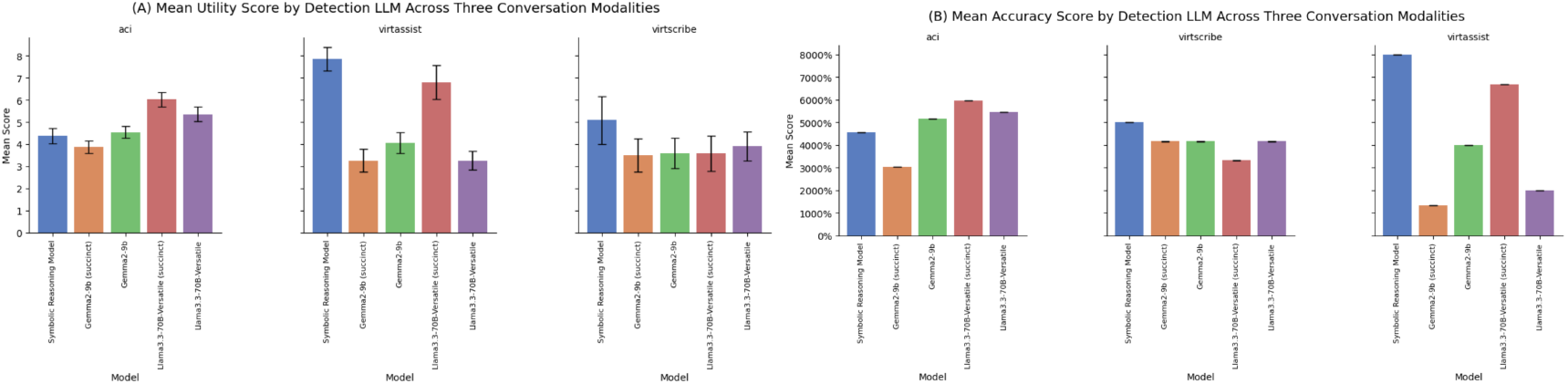
(A; left) Average utility [1–10] and (B; right) Average accuracy [T/F], as scored by three physician reviewers, for chief complaints detected by each model under three conversational modalities.

**Table 1.** (A; top) Average +/– SD utility score and (B; bottom) Average +/– SD accuracy score for each model within each conversation subtype, compared with cross-conversation and cross-model averages. Cross-models measure across all models, within the conversation groupings. Cross-conversation measures across each model, within all conversation groupings.

| Model Name | ACI | Virtscribe | Virtassissit | Cross-conversation mean |
| --- | --- | --- | --- | --- |
| Symbolic Reasoning Model | 4.361 ± 0.422 | 5.083 ± 0.382 | 7.867 ± 1.528 | 5.770 ± 0.777 |
| Gemma-9b-succinct | 3.879 ± 1.500 | 3.500 ± 2.136 | 3.267 ± 2.043 | 3.548 ± 1.893 |
| Gemma-9b-verbose | 4.545 ± 1.756 | 3.583 ± 2.765 | 4.067 ± 1.501 | 4.065 ± 2.007 |
| Llama-3.3-70b-succinct | 6.048 ± 1.149 | 3.583 ± 2.402 | 6.800 ± 0.346 | 5.477 ± 1.299 |
| Llama-3.3-70b-Verbose | 5.364 ± 1.553 | 3.917 ± 2.504 | 3.267 ± 1.286 | 4.182 ± 1.781 |
| Cross Model Mean | 4.839 ± 1.276 | 3.933 ± 2.038 | 5.053 ± 1.341 | 4.609 ± 1.552 |
| Symbolic Reasoning Model | 45.5 ± 15.7% | 50.0 ± 0.00% | 80.0 ± 34.5% | 58.5 ± 16.8% |
| Gemma-9b-succinct | 30.3 ± 22.9% | 41.6 ± 38.2% | 13.3 ± 11.5% | 28.4 ± 24.2% |
| Gemma-9b-verbose | 51.5 ± 29.2% | 41.6 ± 38.2% | 40.0 ± 20.0% | 44.4 ± 29.0% |
| Llama-3.3-70b-succinct | 59.6 ± 20.3% | 33.3 ± 38.2% | 66.7 ± 11.5% | 53.2 ± 23.3% |
| Llama-3.3-70b-Verbose | 54.5 ± 24.0% | 41.6 ± 38.2% | 20.0 ± 20.0% | 38.7 ± 27.4% |
| Cross Model Mean | 48.3 ± 22.4% | 41.6 ± 30.5% | 44.0 ± 19.5% | 44.7 ± 24.2% |

The combination of conversation modality and chief complaint detection model that produced the best output were the SRM in virtassist conversations, which achieved an average utility score of 7.867 and average accuracy of 80% (Table 1). The SRM also achieved the highest scoring detections in virtscribe (5.083 average utility, 50% average accuracy) conversions compared to Llama and Gemma. However, the SRM records the second lowest clinical utility score in the aci conversation modality (4.361 utility, 45.5% accuracy). Across conversation modalities, the Gemma models prompted for succinctness consistently perform worse than when they are prompted without restrictions. In virtscribe and aci conversations, succinct Llama seemingly achieves greater utility (virtscribe: 6.800 v.s 3.267; aci: 6.048 v.s 5.364), but this effect is reversed in virtassist conversations (3.583 v.s 3.917).

The utility metrics of each model were collected by running the models multiple times (n_SRM_=200, n_LLMs_=30) to predict across the test set and averaging across all iterations. The SRM has a significantly lower run time than the LLM models, but with a significantly higher computational cost on the local RAM memory.

## Discussion

### Key Findings

In this investigation, we sought to compare the performance of out-of-the-box LLMs against a rules-based SRM at chief complaint detection in clinical conversations. The SRM leveraged Zipf’s Law to capture linguistic patterns and conversational behavior commonly identified in patient-physician communication. We evaluated each model on simulated test set dialogues acted by real physicians and patient actors in the Aci-bench dataset^[7]^.

The SRM recorded best-in-class performance on the virtscribe and virtassist conversation modalities by using conversational flags that occur commonly in certain medical dialogues (i.e “patient presents with…”). The linguistic patterns of virtscribe and virtassist compared to the aci conversations contribute to the enhanced performance of the SRM in non-aci conversation modalities, due to the conversational flags such as “patient is here for…” or “patient presents with…”. Although the SRM ranks fourth against LLMs in chief complaint detection on aci conversations, it remains competitive with Gemma2-9b, prompted with and without succinctness. Thematic reasoning, or the ability to form inferences from recurring topics, ideas, and patterns without prompting, is a unique strength of larger LLMs, such as Llama3.3-70B-Versatile. However, this discerning thematic reasoning capability comes at a computational cost tradeoff. Llama3.3-70B-Versatile takes significantly more time than the SRM and has the additional cost per token (Supplementary table 2). It is worth noting that Gemma2-9b has lower costs to Llama3.3-70B-Versatile, but with higher runtimes and worse utility scores.

Furthermore, although the locally run SRM uses 40 times more RAM compared to LLMs, it is worth acknowledging that cloud-based LLMs were evaluated through API calls which dramatically reduce their RAM consumption. Deploying LLMs involves additional resource burdens, such as significant cloud infrastructure costs, which can range from $3–4 USD/month per user for a 70b model in dedicated environments^[10]^ and specialized staff to effectively manage these softwares.

### Limitations

This study has a small sample size for conversational data input. Further development of a SRM within hospital systems who have larger datasets of doctor-patient conversations is required for a model that is clinic-ready. We also acknowledge this study ran LLMs only thirty times in Figure 3. These LLMs still demonstrate significantly greater runtime than the SRM. These reasons limit our findings to a proof-of-concept that an SRM is a comparable option to LLMs for resource-limited healthcare settings.

**Figure 3.**
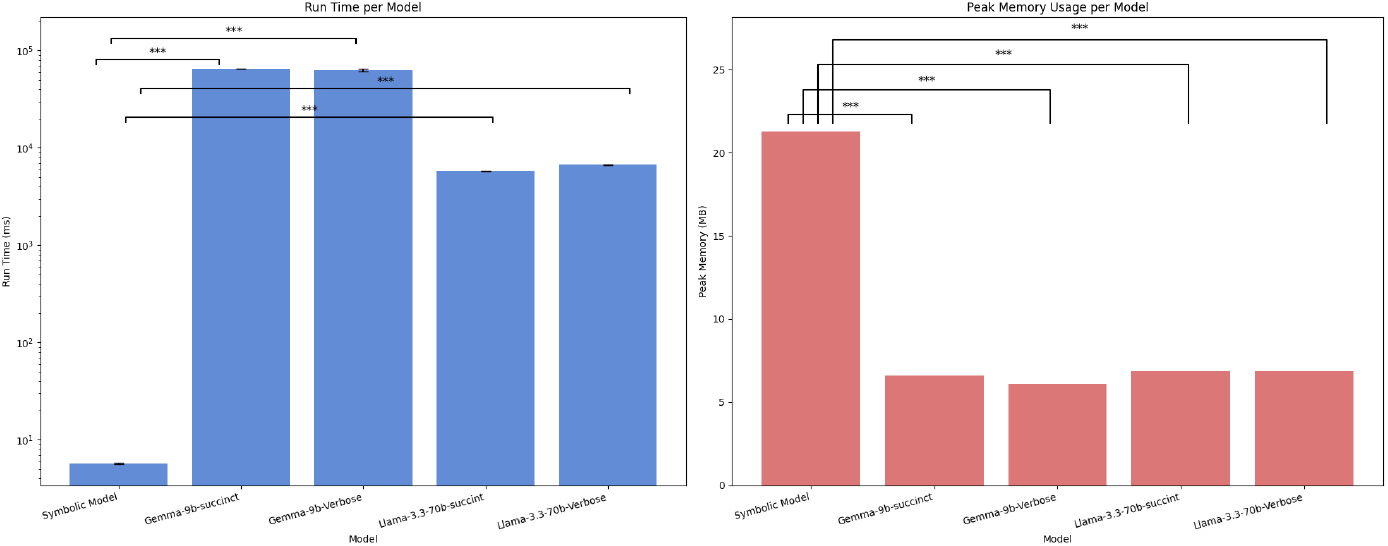
Run time and memory usage per model. Symbolic n=200, LLM models n=30. Run time (blue) in ms to run the models and RAM memory usage (yellow) in MB to run the models. LLMs were API calls, while the symbolic model was run locally.

### Future Directions

Our findings indicate that our SRM can match the performance of LLMs, such as Gemma2-9B and Llama3.3-70B-versatile at the detection of chief complaints from the synthesized dataset. Future work can evaluate the performance of SRMs on larger datasets with more diverse conversational settings. For instance, comparing the performance of these models on transcriptions with regional differences in language or between different subspecialties of outpatient care may reveal inherent strengths and/or limitations of SRMs for virtual clinical scribing. Finally, developing SRMs to detect more than chief complaints (i.e. lab summaries, patient history, etc.) from doctor-patient conversations would bridge the gap in applying SRMs to clinical practice.

## Conclusions

We demonstrate here a preliminary proof-of-concept that, at a shorter runtime and lower operational cost, an SRM outperforms state-of-the-art LLMs at chief complaint detection with minor conversational adjustments from a physician to acknowledge a third-party scribe system. This system could resolve bottlenecks in outpatient workflows and reduce medical documentation costs for healthcare systems that lack the resources or funding to safely implement LLMs, such as smaller hospitals or clinics in rural settings^[5,11]^. While it may take time to transition to AI and generative AI applications, organizations seeking to automate clinician summaries and reduce physician burnout can begin by employing SRMs.

## Clinical Relevance Statement

Effective clinical scribing is a meticulous, expensive, and time-consuming task in medical care. The findings of this study support the development of algorithmic digital scribes to address this unresolved challenge at a lower operational cost than LLM and generative AI-powered scribe assistants. When physicians use conversational cues designed to trigger a scribe assistant, an algorithmic SRM detects patient chief complaints better than LLMs, as scored by three physicians from different practices. This finding has potential to be integrated into low-cost digital scribe systems, mobile health assistants, and improve access to ambient clinical intelligence systems in under-resourced healthcare settings.

## Data Availability

All data produced in the present study are available in Supplementary Table 1. All scripts to reproduce these analyses are available online at https://github.com/david-donofrio/datathon.

## Acknowledgements

The authors would like to thank the three physicians who scored the accuracy and utility of detected chief complaints, as well as the leadership from the non-profit organization MD+, whose feedback spurred iterations on the models and benchmarks presented here.

## Conflict of Interest

None declared.

## Protection of Human and Animal Subjects

No patient data or animal subjects were used in this study. Simulated conversations between real physicians with actor patients were anonymized, transcribed, and published by Microsoft Corporation.

## Notes

### Competing Interest Statement

The authors have declared no competing interest.

### Funding Statement

This study did not receive any funding.

