## Supplementary for "Extracting clinical chief complaints from patient-physician conversations with a symbolic reasoning model"

**Supplemental Material**

**Supplementary Table 1:** SRM, Gemma-9b, and Llama-3.3-70b extracted chief complaints and their corresponding physician utility scores and accuracy grading for all test set conversations in aci-bench.

(Attached separately)

**Supplementary Table 2:** Usage metrics across all models for run time, local RAM usage, and token cost. These models were run N times and average scores and standard deviations across each metric were taken, when applicable. The SRM does not have a cost associated because it is run locally and does not require API calls like the LLM models.

| **Model** | **Symbolic Reasoning Model** | **Gemma-9b-Succinct** | **Gemma-9b-Verbose** | **Llama-3.3-70b-Succinct** | **Llama-3.3-70b-Verbose** |
|---|---|---|---|---|---|
| **N Runs** | 200 | 30 | 30 | 30 | 30 |
| **Time Average ± SD (ms)** | 5.73 ± 0.10 | 65408.52 ± 75.63 | 63359.42 ± 1976.70 | 5777.58 ± 48.26 | 6720.84 ± 58.76 |
| **Maximum local RAM usage (MB)** | 21.30 | 6.63 | 6.11 | 6.88 | 6.88 |
| **Token Cost Average ± SD ($)** | N/A | 0.0065 ± 0.0000018 | 0.0065 ± 0.0000020 | 0.019 ± 0.00000059 | 0.019 ± 0.0000079 |

**Supplementary Figure 1:** Spearman's rank correlation coefficient calculated between physician reviewer utility scores and word count length of chief complaints. Reviewers are generally in agreement with what complaints deserve higher versus lower utility scores. There is no preferential bias toward longer or shorter chief complaints.

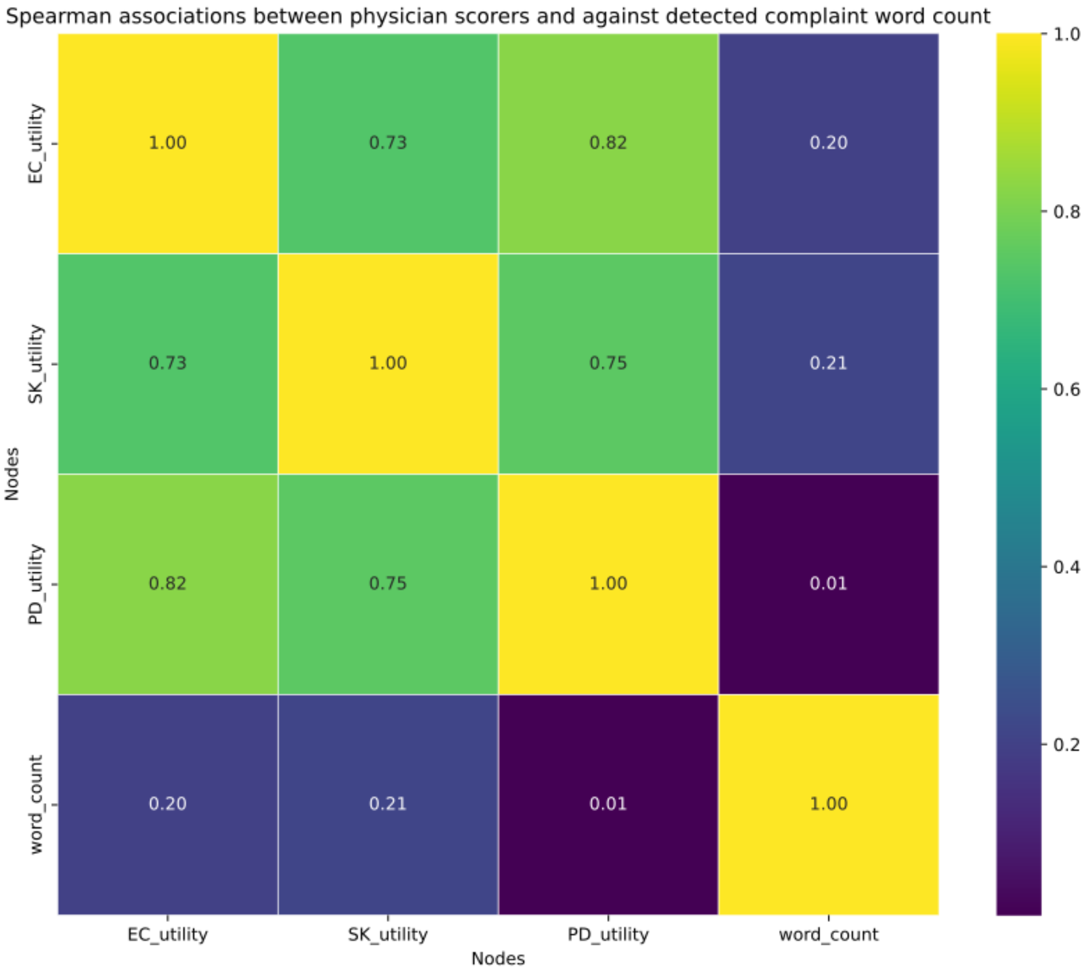

**Supplementary Figure 2:** Physician reviewer concordance across conversation types. Reviewer EC and PD agree on the accuracy (T/F) of 49/55 aci conversation chief complaints, 22/25 in virtassist conversation chief complaints, and 15/20 virtscribe chief complaints. All three reviewers agree on chief complaint accuracy on 32/55 aci, 16/25 virtassist, and 8/20 virtscribe conversation chief complaints.

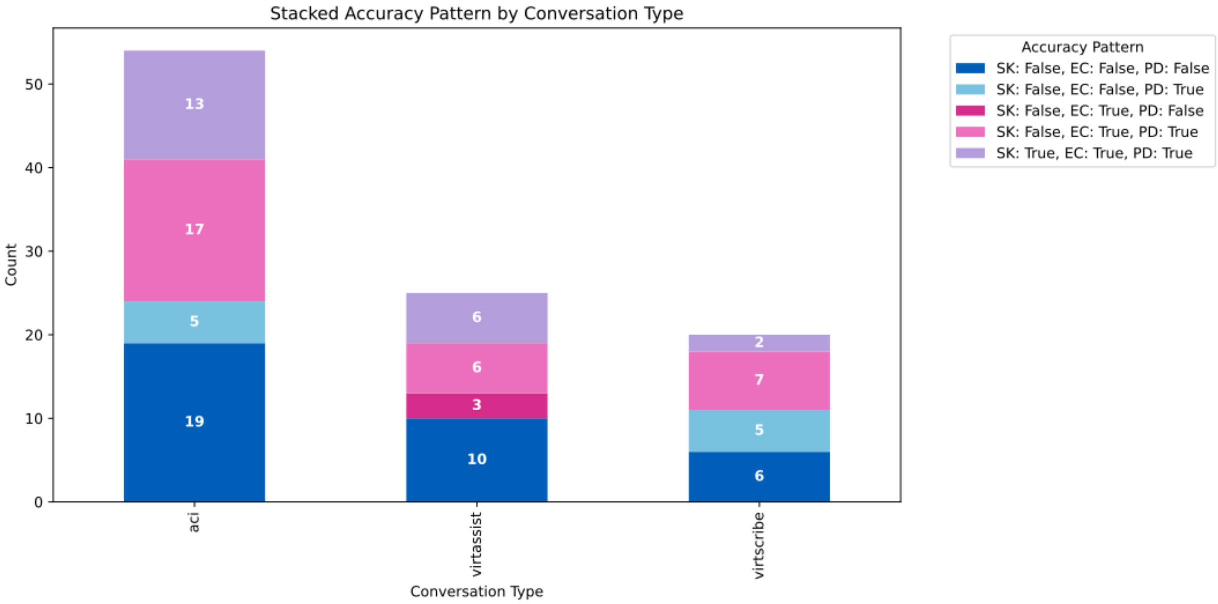
